# Elevation of cell-associated HIV-1 RNA transcripts in CSF CD4+ T cells, despite suppressive antiretroviral therapy, is linked to in vivo brain injury

**DOI:** 10.1101/2021.12.22.21268288

**Authors:** Kazuo Suzuki, John Zaunders, Thomas M. Gates, Angelique Levert, Shannen Butterly, Zhixin Liu, Takaomi Ishida, Chin-Shiou Huang, Sarah Palmer, Carolin D. Rae, Lauriane Jugé, Lucette A. Cysique, Bruce J. Brew

## Abstract

**Objective:** Despite effective antiretroviral therapy (ART), brain injury remains prevalent in people living with HIV-1 infection (PLHIV) possibly due to ART’s lack of direct inhibition of transcription with continued local production of viral transcripts and neurotoxic proteins, such as Tat, rather than cell-free whole virion toxicity. We quantified cell-associated (CA) HIV-1 RNA-transcripts in CSF and blood, in relation to proton Magnetic Resonance Spectroscopy (^1^H MRS) of major brain metabolites, in well characterised PLHIV.

**Methods:** RNA was extracted from cells in 16 paired samples of CSF and blood, from PLHIV on fully suppressive ART. HIV-1 CA-RNA copies were measured using the highly sensitive Double-R assay and normalized /10^6^ CD4+ T cells. 18-colour flow cytometry was used to count and analyse CD4+ T cells and monocytes in CSF and blood. The concentrations of major brain metabolites from ^1^H MRS in frontal white matter (FWM), posterior cingulate cortex (PCC), and caudate areas were measured. Brain injury in each voxel was defined using a composite score derived by principal component analysis.

**Results:** 14/16 CSF cell samples had quantifiable HIV-1 CA-RNA transcripts, at levels significantly higher than in their PBMCs (median 9,266 vs 185 copies /10^6^ CD4+ T cells; p<0.0001). Higher levels of CSF transcripts were associated with greater brain injury in the FWM (Std β=-0.73; p=0.007) and PCC (Std β=-0.61; p=0.03). CSF cells were 91% memory T cells, equally CD4+ (median 3,605) cells and CD8+ T cells (3,632), but contained much fewer B cells (0.4 %), NK cells (2.0%) and monocytes (3.1%; 378 cells; >90% CD14+CD16+ phenotype). CXCR3+CD49d+integrin ß7-negative, CCR5+ CD4+ T cells were significantly enriched in CSF, compared with PBMC (p <0.001). Transcriptional activity in CSF cells was highly correlated with levels of transcriptional activity in CD4+ T cells in PBMC (r=0.76; p=0.002). In contrast, HIV-1 RNA in highly purified monocytes from PBMC was detected in only 6/16 samples.

**Conclusions:** Elevated HIV-1 transcripts in CSF cells were associated with in vivo brain injury, despite suppressive ART. The cellular source is most likely the predominant CXCR3+ CD49d+ integrinß7-CCR5+ memory CD4+ T cells, not monocytes. Inhibitors of transcription to reduce local production of potentially neurotoxic proteins, should be developed.

## INTRODUCTION

Despite suppressive antiretroviral therapy (ART) leading to undetectable HIV-1 RNA in both plasma and cerebrospinal fluid (CSF), in vivo brain injury in people living with chronic HIV-1 infection (PLHIV), revealed by proton magnetic resonance spectroscopy (^1^H MRS), persists and remains common (1).

The mechanisms underlying this persistent brain injury remain unclear. Indeed, in chronic HIV-1 infection, concurrent mechanisms of brain injury may include neurological and psychiatric confounds, age-related comorbidities, legacy effect of pre-ART deficit, poor brain penetration of ART, but also possible ART toxicities (2, 3). Nonetheless, a direct HIV-1 cause of brain injury in chronic HIV-1 infection cannot be excluded when the role of viral reservoirs and sanctuaries is considered, especially viral replication within the central nervous system (CNS). Consideration of brain injury despite suppressive ART relates to the reservoir and sanctuary size (HIV-1 DNA copy number), the location within or outside the CNS, the transcriptional activity (cell-associated (CA) HIV-1 RNA copy numbers) and the virus replication competence (cell-free virus). The current interpretation of the reservoir and sanctuary as they pertain to the CSF (as an indirect marker of the brain) has been influenced by assay methods that have variably detected HIV-1 DNA or RNA (4, 5).

The Double R assay, based on the πCode End-Point PCR platform is much more sensitive than previous assays at detecting HIV-1 DNA and RNA (6, 7). It can detect both spliced and unspliced mRNA which can then be translated into viral proteins such as Tat, Env, Nef, Rev, and Vpr. These are important as they are neurotoxic and can still be produced from the reservoir even though most of the reservoir cannot produce replication competent whole virus (8, 9). Furthermore, suppressive ART does not stop production of these viral components once HIV-1 is integrated into the host genome; ART targets almost every step of the life cycle of HIV-1 from receptor-mediated entry to budding from the cell surface. However, it does not specifically affect transcription of RNA from the promoter in the integrated HIV-1 proviral DNA. Therefore, transcription and then translation, of early viral proteins such as Tat may still occur despite suppressive ART (3). We therefore hypothesized that brain injury is related to the production of these proteins in the context of suppressive ART.

We tested this hypothesis by using the highly sensitive Double R assay to detect and quantify HIV-1 DNA copy number and CA-RNA transcripts in CSF cells and PBMCs from PLHIV on suppressive ART, correlating the findings with brain injury as assessed by ^1^H MRS. We also examined the cellular composition of the CSF to determine the likely source of these transcripts.

## METHODS

### Participants

The 16 study participants (Table S1) were HIV-1 infected males on fully suppressive ART (as assessed in both blood and CSF) who were enrolled into an ongoing prospective study of CNS HIV-1 latency and NeuroHIV biomarkers (ClinicalTrials.gov Identifier: NCT02989285). To be included in this current study, participants were required to have had stable HIV infection with viral suppression for at least 6 months, and sufficient fresh blood and CSF had to be available.

The study protocol was approved by the St. Vincent’s Hospital Human Research Ethics Committee (HREC/15/SVH/425) and all participants provided written informed consent prior to enrolment.

### Collection and processing of CSF cells and peripheral blood mononuclear cells (PBMC)

CSF and anti-coagulated peripheral blood were collected via lumbar puncture and phlebotomy, respectively, during one of the five study visits (a screening/baseline visit and four 6-monthly follow-up visits over 24 months).

CSF samples (median 12.6 ml; IQR: 10.4-13.7 ml) were centrifuged at 400g for 15 min. CSF was carefully aspirated and the cell pellets were re-suspended in 1ml Dulbecco’s Ca^2+^ and Mg^2+^ free PBS (Gibco, Life Technologies, Paisley, UK) containing 2% fetal calf serum (FCS; Gibco).

A 100µl aliquot of CSF cell suspensions was analyzed by Flow Cytometry (see below), and the remaining 900µl of CSF cell suspensions were centrifuged at 5000g for 3 min to obtain the CSF cell pellets, which were then subjected to extraction of total nucleic acid for HIV-1 molecular analysis by the Double R assay (described below).

Acid-citrate (ACD) anti-coagulated blood samples were obtained on the same date as the Lumbar puncture. PBMCs were prepared from the blood samples by standard density gradient centrifugation on Ficoll-Hypaque Plus (GE Healthcare, Chicago, IL, USA). PBMC were cryopreserved in heat inactivated, filter sterilized, FCS (Gibco) containing 10% dimethyl sulphoxide (DMSO; Sigma Aldrich, MO, USA) using a controlled rate freezer (Planer, Middlesex, UK) and stored in vapour phase liquid nitrogen.

### The Double R assay on the πCode End-Point PCR platform

We used our recently described assay to detect HIV-1 CA-RNA transcription and total HIV-1 DNA copies (6, 7). Briefly, the primers and probes used in this assay target the highly conserved “R” region in both the 5′- and 3′-“LTR” regions, which permits detection of total spliced and unspliced mRNA transcripts, using reverse transcription, as well as, in parallel, integrated HIV-1 proviral DNA without RT (6, 7). The amplicons are then read out with the highly sensitive image analysis of precision image pi-code (nCode) MicroDiscs, as previously described (6, 7). The overall method results in at least 27-fold higher sensitivity than current Real-Time PCR assays (6, 7).

For PBMCs, HIV-1 CA-DNA and RNA were extracted using the Maxwell RSC automated extraction platform (Promega), with the Maxwell RSC Buffy Coat DNA kit (Cat No. AS1540, Promega) and Maxwell RSC Simply RNA Tissue kit (Cat No. AS1340, Promega), respectively, according to the manufacturer’s protocol. The RNA assay for PBMC’s uses the PrimeScript One step RT-PCR kit (Takara Bio, Kusatsu, Shiga, Japan). The Double R assay can detect as few as two OM10.1 cells (containing single integrated HIV-1 provirus per cell) when diluted in as many as 10^6^ uninfected cells (6) (see Supplementary Method for detailed analysis and Supplementary Table 6-8). Quantification of HIV-1 copy number per patient sample was determined using set-4 and set-6 probes (6, 7) with a standard curve generated with HIV-1 plasmid controls: 0.73, 2.2, 6.6, 20, 59, 177, 533 and 1,600 HIV-1 copies /µl. The HIV-1 copy number for each sample, from the average value of duplicate reactions, was normalized with the CD4+ T cell count number, which was obtained by the flow cytometry analysis (below). HIV-1 copy number per one million CD4+ T cells was used as the standardized unit.

CSF cell nucleic acids were obtained using Total Nucleic Acid extraction (Maxwell RSC viral Total Nucleic Acid Purification kit (Cat No. AS1330, Promega, Madison, WI, USA). Total HIV-1 RNA and HIV-1 DNA copy numbers were measured using the PrimeScript One step RT-PCR kit, as above for PBMC. In parallel, HIV-1 DNA copy numbers were identified using the same detection procedure of the PrimeScript One step RT-PCR kit (Takara) without addition of RT enzyme. HIV-1 RNA copy numbers were therefore calculated by subtracting the HIV-1 DNA copy number from the total HIV-1 copy number (see Supplementary Method for detailed validation of this analysis and Supplementary Table 9).

For PBMCs, HTV-1 CA-DNA and RNA were extracted using the Maxwell RSC automated extraction platform (Promega), with the Maxwell RSC Buffy Coat DNA kit (Cat No. AS1540, Promega) and Maxwell RSC Simply RNA Tissue kit (Cat No. AS1340, Promega), respectively, according to the manufacturer’s protocol.

From PBMCs, purified CD14+ monocyte and CD14-negative preparations (see below), respectively, were divided into two portions each with equal numbers of cells, followed by centrifugation at 5000g for 5 min. One cell pellet was used for DNA extraction and the other cell pellet was used for RNA extraction (as for unfractionated PBMC, above).

### Detection of cell-free HIV-1 in CSF

CSF cell-free virus was measured using the standard Roche Amplicor assay with a lower limit of detection of <80 copies/ml, and also using 7 ml of CSF fluid fraction (after centrifugation of cells), in triplicate, on the single copy HTV-1 RNA assay, as previously described (10), with a lower limit of detection of 0.3 copies/ml.

### CD14+ monocyte isolations from PBMC

CD14+ monocytes were isolated from cryopreserved PBMCs, which were thawed and resuspended in PBS with 2% FCS, using positive selection on the automated RoboSep platform (CD14+ cell positive separation kit, Cat No. 17858RF, StemCell Technology, Vancouver, Canada), according to the manufacturer’s directions. The number of CD4+ T cells in the isolated CD14+ monocyte populations and in the CD14-negative fraction were accurately counted using TruCount tubes (BD Biosciences). CD14+ monocyte fractions were routinely highly purified and only contained median 0.26% (TQR: 0.19-0.35) contaminating CD4 T cells.

### Flow cytometry analysis

As described above, CSF samples were centrifuged at 400g for 15 min. CSF was carefully aspirated and the cell pellet resuspended in 1ml Dulbecco’s Ca^2+^ and Mg^2+^ free PBS containing 2% FCS. A 40 µl aliquot of these CSF cells was used to accurately count CD4+ T cells and other PBMC types, using a 12-colour monoclonal antibody panel (Table S2a), in TruCount tubes (BD Biosciences, San Jose, CA, USA), according to the manufacturer’s directions, on a 5-laser Fortessa flow cytometer (BD Biosciences), as previously described (11). A 60 µl aliquot was separately further analyzed for memory T cell subsets using an 18-colour monoclonal antibody panel (Table S2b), on the Fortessa flow cytometer as previously described (12). Cryopreserved PBMC samples were thawed and analysed for memory T cell subsets using the same 18-colour monoclonal antibody panel (Table S2b).

### ^1^H MRS

^1^H MRS brain scans were conducted within 4 weeks of CSF and PBMC collection for the majority (60%) of participants (median 0 weeks; TQR: -1-62 weeks). The ^1^H-MRS protocol used in this study has been described previously (13). Briefly, spectra were acquired on a Phillips 3T Tngenia scanner (Philips, Best, Netherlands) using 32-channel head coil. Cerebral metabolite concentrations were quantified using point-resolved spectroscopy (PRESS; TE=40ms; TR=2000ms) in the frontal white matter (FWM), posterior cingulate cortex (PCC), and caudate nucleus. jMRUT v3.0 (14) with AMARES algorithm (15) was used to analyse fitted spectra, which included *N*-acetyl aspartate (NAA), choline (Cho), creatine (Cr), *myo*-inositol (mTo), and glutamate (Glu). Spectra were expressed as ratios in relation to unsuppressed water signal (H_2_O). Spectra values were normalized using a reference sample of demographically comparable 54 HTV-controls from our HTV-1 and Ageing Research Program (16) and converted to age-corrected z-scores.

Age-corrected spectra z-scores were highly correlated within each voxel (FWM: ρs=.20-.84; PCC: ρs=.56-.89; caudate ρs=-.02-.66). The metabolites’ data by voxel were amenable to data reduction using principal components analysis and maximum likelihood method, a method of choice to reduce ^1^H MRS data (17). We extracted a single in vivo composite score for each voxel accounting for 67% of common variance in the FWM, 84% in the PCC, and 71% in the caudate spectra (see Supplementary Figure 1 for relative contributions of each metabolite to voxel composite scores). Lower ^1^H MRS composite scores reflected higher levels of in vivo brain injury.

### Neuropsychological Methods

We used a standard neuropsychological battery (see Supplementary Methods and Supplementary Table 5) covering 7 cognitive domains, in line with the Frascati recommendations (18) and NeuroHTV research internationally (19), that has previously been demonstrated by our group to be sensitive to HAND (20, 21).

### Statistical analysis

Standard curves from known concentrations of HIV-1 plasmid copy numbers were generated with GraphPad Prism v7 (GraphPad Software). HIV-1 CA-RNA transcription and HIV-1 CA-DNA copy numbers (as measured by the Double R assay) in CSF cells vs PBMCs were compared by non-parametric Mann-Whitney U tests. Differences in the proportions of subsets of memory CD4+ T cell subsets, between paired CSF cell and PBMC samples, from individual participants, were compared using paired Wilcoxon non-parametric tests. Pearson correlations were used to analyze the association between HIV-1 CA-RNA and HIV-1 CA-DNA levels within each cell type.

To investigate the impact of suppressive ART on transcriptional activity we divided the total duration of HIV-1 infection into two time-related variables that were examined separately: time from seroconversion to suppressive ART initiation, and duration of suppressive ART exposure. Non-parametric Spearman’s rank correlations were used to analyze the relationships of HIV-1 CA-RNA transcription and HIV-1 CA-DNA levels in CSF and PBMCs with suppressive ART exposure. We additionally classified participants as being either “early” or “late treated”, depending on whether or not they commenced suppressive ART within vs more than 12 months post-seroconversion.

To characterize the association between HIV-1 CA-RNA transcription and HIV-1 CA-DNA copies with ^1^H MRS-related brain inJury, we conducted a series of hierarchical linear regression models. Each model contained one of the four Double R assay biomarkers (i.e., CSF CD4+ HIV-1 CA-RNA transcription, CSF CD4+ HIV-1 CA-DNA, PBMC CD4+ HIV-1 CA-RNA transcription, or PBMC CD4+ HIV-1 CA-DNA) as the criterion variable. In the first step, one of the three ^1^H MRS voxel composite scores was entered. In the second step, early vs late suppressive ART initiation was entered as a covariate. We also considered models where time to suppressive ART initiation was entered instead as a continuous covariate at step 2. However, we retained the models containing the dichotomous suppressive ART exposure variable as it provided a similar or slightly improved fit each time. Parameter estimates and standardized β effect sizes were extracted at each step of model fitting and compared.

For two participants, two sets of PBMC/CSF cell pairs were collected six months apart. We used these additional samples to assess the longitudinal stability of the molecular-based analyses.

## RESULTS

### Sample characteristics

The cohort comprised 16 PLHIV (Table S1). They were aged 63.5 years on average, English speaking background Australian white men with chronic, treated HIV (median HIV infection duration 28 years, on suppressive ART for a median 22 years, ten (62.5%) initiated suppressive ART more than 12 months post-seroconversion). Ten (62.5%) had an historical diagnosis of AIDS. The sample was otherwise homogeneous in terms of their global cognitive functioning which was close to or slightly below the normative mean, reflecting some degree of chronic vulnerability. Six participants (37.5%) had an historical diagnosis of HIV-associated neurocognitive disorder (HAND) and four (25%) had current mild HAND (1 ANI, 3 MND). Median nadir CD4+ T cell count was 167 cells/µl and current median CD4+ T cell count was 721.5 cells/µl.

### Detection of cell-free HIV-1 in CSF and plasma

CSF cell-free HIV RNA was <80 copies/ml for all samples, using the Roche Amplicor assay, and furthermore, the single copy assay of cell-free HIV-1 RNA in CSF was undetectable in each of the 13 (81%) CSF samples with sufficient volume available for testing.

Plasma samples were fully virally suppressed with HIV RNA <50 copies/ml using the Roche Amplicor assay. Single copy assay of HIV-1 RNA in plasma was undetectable in eight (50%) samples (<0.3 copies/ml). In the remaining eight detectable samples, the median was 37 copies/ml.

### Detection of cell-associated (CA) HIV-1 in CSF cells and PBMCs using the Double R assay

The cells in the CSF were dominated by CD3+ T cells, which were evenly divided between CD4+ and CD8+ T cells (Supplementary Figure 1a). Other cell types in the CSF were proportionally much lower, including monocytes (3.1% of CSF cells; Supplementary Figure 1b), NK cells (2.0%) and B cells (0.4%). The numbers of these cells recovered in the CSF samples are shown in Figure 1a. In particular, the number of CD4+ T cells (median: 3,605 cells) greatly outnumbered monocytes (378 cells), such that, in all but two samples, there was an average >20-fold difference. Furthermore, the monocytes were >90% CD14+CD16+, the phenotype of intermediate monocytes in blood.

**Figure 1.**
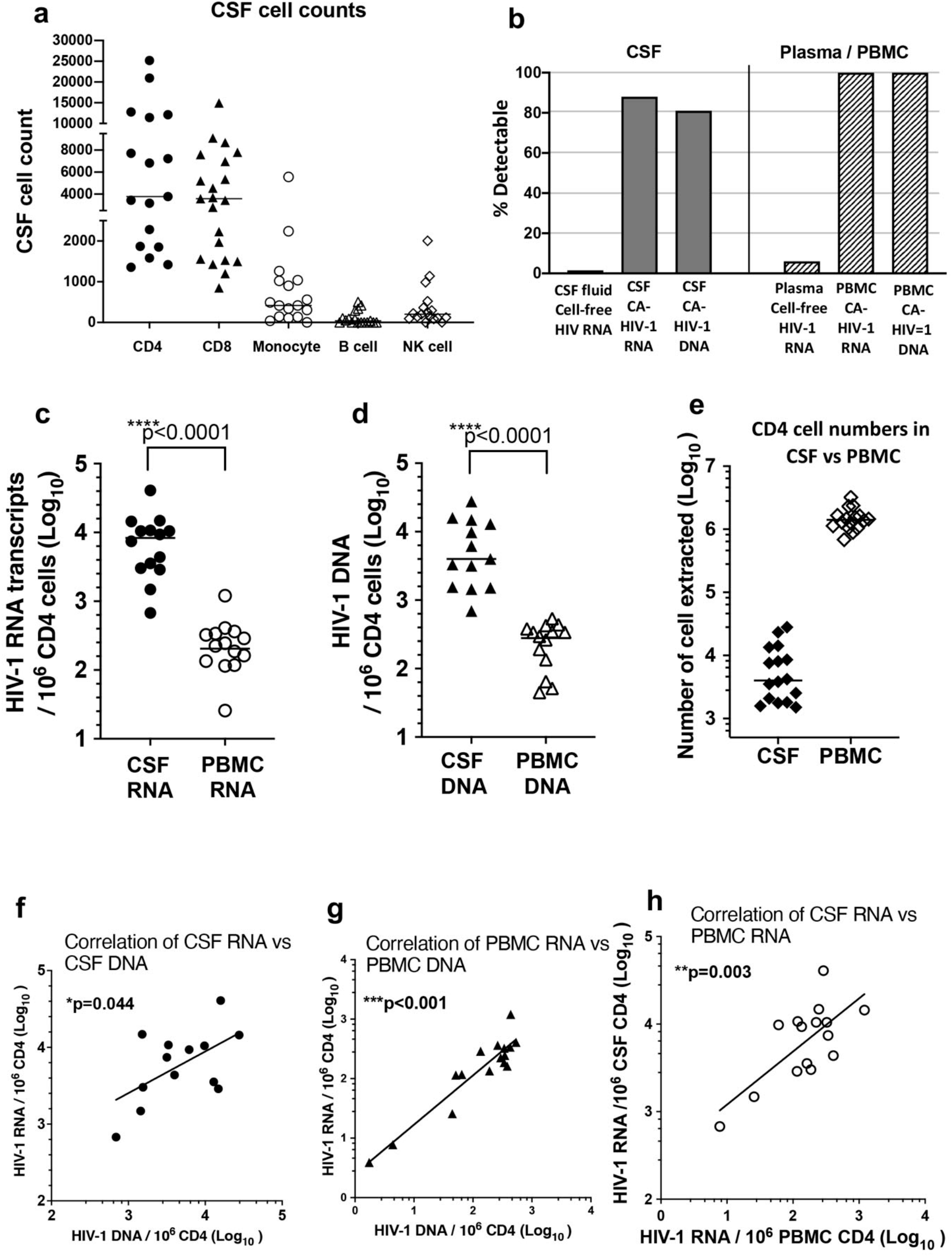
Comparison of CSF and PBMC cell numbers and HIV-1 CA-RNA and DNA levels. (**a**) Range of recovered CSF cell counts for different CD45+ cells obtained in the pellets, including CD3+CD4+ and CD3+CD8+ T lymphocytes, CD14+ monocytes, CD19+ B cells and CD16+CD56+ NK cells; (**b**) Rate of detectability of cell free virus using single copy assay, in CSF and plasma, respectively and cell-associated (CA) HIV-1 RNA and DNA in CSF cells and PBMC, respectively; (**c**) comparison of HIV-1 CA-RNA copies, normalized per 10^6^ CD4 T cells, for CSF cells and PBMC, respectively; (**d**) comparison of HIV-1 CA-DNA copies, normalized per 10^6^ CD4 T cells, for CSF cells and PBMC, respectively; (**e**) numbers of CSF cells and PBMC used for extractions; (**f**) correlation of copy numbers of HIV-1 CA-RNA vs DNA in CSF cells; (**g**) correlation of copy numbers of HIV-1 CA-RNA vs DNA in CSF cells; and (**h**) correlation of copy numbers of HIV-1 CA-RNA in CSF cells vs HIV-1 CA-RNA in PBMC.

These CSF cells contained detectable HIV-1 CA-RNA transcription copies in 14/16 (88%) participant samples and HIV-1 CA-DNA copies were detected in CSF cells from 13/16 (81%) participant samples (Figure 1b, left). This contrasted with the undetectability of cell-free HIV-1 RNA in the fluid fraction, using either the diagnostic Amplicor or single copy HIV-1 RNA assays (see above).

In PBMCs, HIV-1 CA-RNA transcripts and HIV-1 CA-DNA copies were both detected in 16/16 (100%) participant samples (Figure 1b, right). Once again, this contrasted with limited detection of cell-free HIV-1 RNA in plasma, as described above.

The quantitation from the Double R assay was normalized as HIV-1 copy number per 1 × 10^6^ CD4+ T cells, to permit comparison with the corresponding PBMC samples for each participant. The results show that HIV-1 CA-RNA transcription levels were significantly higher in CSF cells than paired PBMCs (Figure 1c: median 9,266 vs 185 copies/10^6^ CD4+ T cells; p<0.0001). Total HIV-1 CA-DNA levels were also significantly higher in CSF cells than corresponding PBMCs (Figure 1d: median 4,021 vs 261 copies/10^6^ CD4+ T cells; p<0.0001). Note that there were 300-fold fewer CD4+ T cells extracted from CSF cells compared to PBMCs (Figure 1e), demonstrating how enriched the HIV-1 CA-RNA transcription and HIV-1 CA-DNA levels were in CSF cells.

Consistent with our earlier study of PBMCs from fully suppressed patients (7), there was a medium correlation between HIV-1 CA-RNA transcription and HIV-1 CA-DNA in CSF cells (Figure 1e; r=0.58; p=0.04). Similarly, there was a medium to large correlation between HIV-1 CA-RNA transcription and HIV-1 CA-DNA in CD4+ T cells from PBMCs (Figure 1f; r=0.72; p<0.001).

Importantly, there was a large correlation between HIV-1 CA-RNA transcription in CSF cells and HIV-1 CA-RNA transcription in CD4+ T cells from PBMCs (Figure 1g; r=0.83; p=0.003).

### HIV-1 CA-RNA and HIV-1 CA-DNA relationships with brain ^1^H MRS

HIV-1 CA-RNA transcription in CSF CD4+ T cells showed an inverse relationship of large size with the FWM (Std β=-0.73, p=0.007) and medium size with the PCC ^1^H MRS composite scores (Std β=-0.61, p=0.03) (Figure 2a; Supplementary Table 3). The univariate association between HTV-1 CA-RNA transcription and the caudate composite score was weaker (Std β=-0.38; p=0.21), although the relationship was comparable in magnitude to that seen in the FWM and PCC once the effect of early vs late initiation of suppressive ART was controlled (Std β=-0.57, p=0.04).

**Figure 2.**
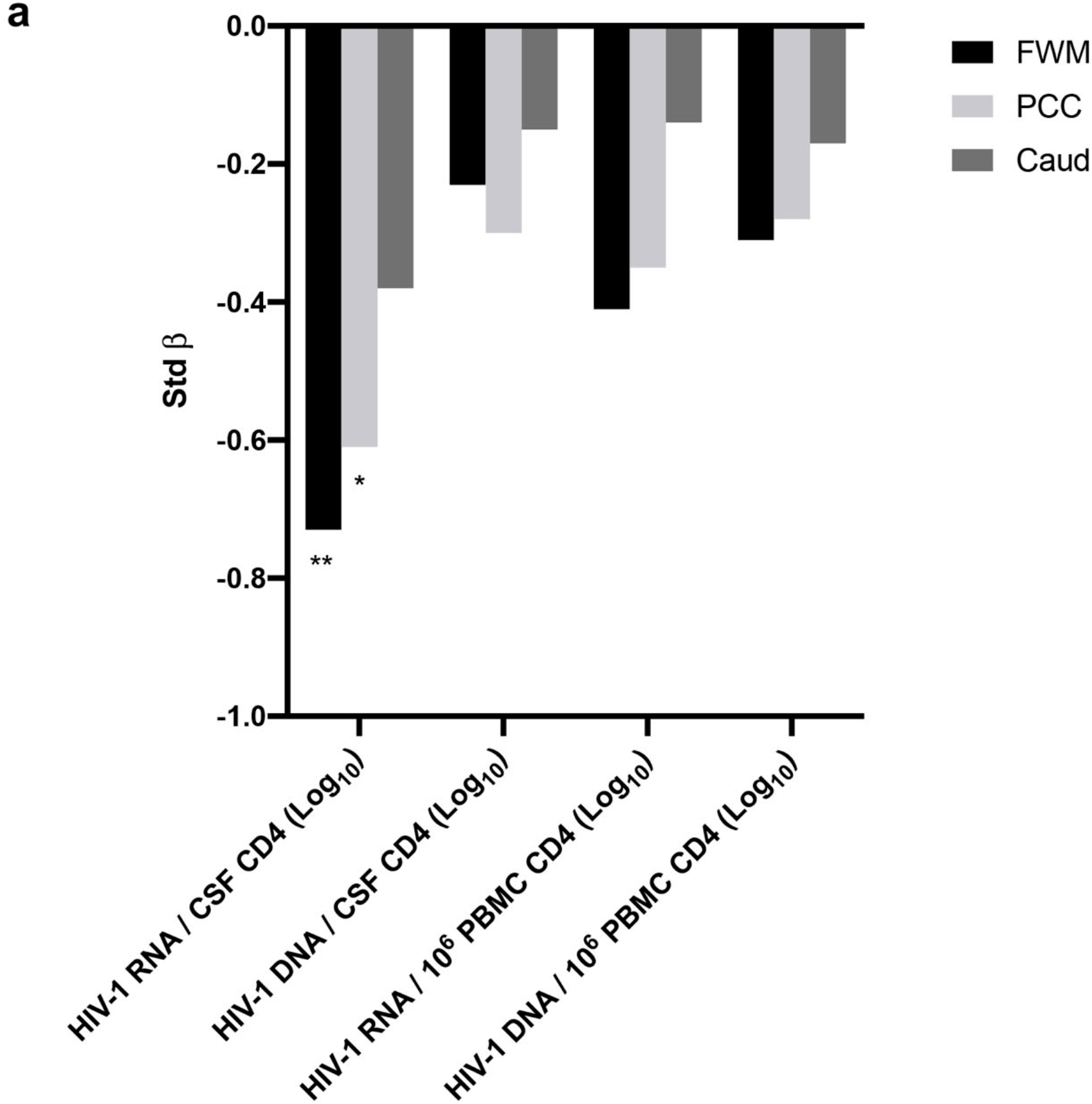
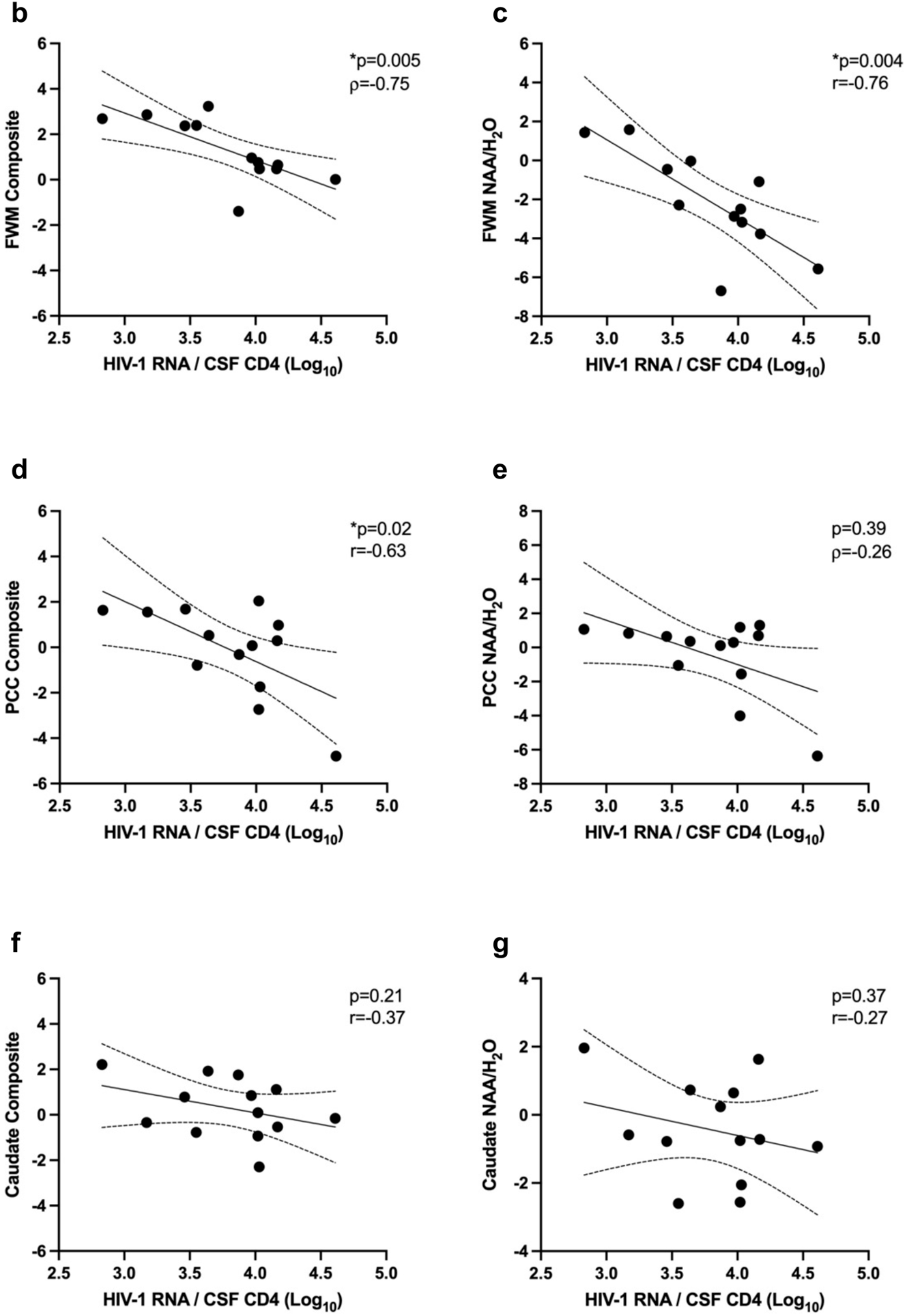
Associations of ^1^H MRS with HIV-1 CA-RNA transcripts and DNA copies. (**a**) HIV-1 CA-RNA and DNA associations with ^l^H MRS voxel composite scores. Univariate associations between log-transformed HIV-1 CA-RNA and DNA copy numbers and age-corrected ^l^H MRS voxel composite z-scores are presented. Asterisk denotes statistical significance at * p<0.05 **p<0.01. For the PCC/CSF HIV-1 CA-RNA model, addition of early vs late initiation of suppressive ART at step 2 led to a small decrease in the strength of the association between the PCC composite score and CSF HIV-1 CA-RNA such that the effect was no longer statistically significant (Std β=-0.53, p=0.06); and correlations of HIV-1 CA-RNA in CSF cells with: (**b**) FWM composite score; **(c)** FWM NAA/H_2_O; **(d)** PCC composite score; **(e)** PCC NAA/H_2_O; **(f)** Caudate composite score; **(g)** Caudate NAA/H_2_O.

The univariate relationships of HTV-1 CA-RNA-transcription levels in PBMC CD4+ T cells with the three ^1^H MRS voxel composites ranged from small to medium in size (Std βs=-0.14 to -0.41; ps>.15). However, when the effect of early vs late initiation of suppressive ART was controlled, the relationship with the FWM composite score became statistically significant (Std β=-0.47; p=0.04) and with the caudate reaching marginal statistical significance (Std β=-0.34; p=0.06).

The associations between HTV-1 CA-DNA levels and the three ^1^H MRS voxel composite scores were non-significant and of small size (in CSF cells: Std βs=-0.15 to -0.30; ps>0.34; in PBMCs: Std βs=-0.17 to -0.31; ps>0.28).

NAA and Creatine contributed the most to the three voxel composite MRS scores (Supplemental Figure 2). We therefore re-conducted the above analysis selecting the NAA/H_2_O ratio (reflecting axonal/neuronal damage) in place of the voxel composite score. Similar to the voxel composite model, a significant large inverse association was observed between FWM NAA/H_2_O and HTV-1 CA-RNA transcription in CSF CD4+ cells (Std β=-0.76; p=0.004) (Supplementary Table 4). There was also a significant medium-sized inverse relationship between FWM NAA/H_2_O and HTV-1 CA-RNA transcription in PBMC CD4+ cells (Std β=-0.76; p=0.004). Tnverse relationships of similar magnitude were also observed between NAA/H_2_O in the PCC and HTV-1 CA-RNA transcription in CSF CD4+ cells (Std β=-0.53; p=0.06), as well as between NAA/H_2_O in the FWM and HTV-1 CA-DNA in PBMC CD4+ T cells (Std β=-0.51; p=0.06), although these effects only reached marginal statistical significance.

### HIV-1 CA-RNA and HIV-1 CA-DNA relationships with time to ART initiation and HAND status

At step 2 of the hierarchical linear regression models, late initiation of suppressive ART (i.e., >12 months post-seroconversion) was consistently associated with HIV-1 CA-RNA transcription and HIV-1 CA-DNA levels in CSF and PBMC CD4+ T cells (Supplementary Table 3). This effect was strongest in the models focused on PBMCs, reaching a medium effect size (Std βs=0.52-0.69; ps<0.05). Similar effect sizes were observed in caudate-based models focusing on CSF HIV-1 CA-RNA and DNA (both Std βs=0.62; ps=0.02-0.06), while smaller (non-significant) effects were observed in FWM- and PCC-based models focused on CSF HIV-1 CA-RNA and DNA (Std βs=0.27-0.47; ps>0.14).

When evaluating suppressive ART exposure as a continuous variable, trends in the same direction emerged, albeit not to a statistically significant level. Specifically, longer time from seroconversion to suppressive ART initiation showed small to medium sized associations with higher HIV-1 CA-RNA-transcription levels (ρ =0.25, p=0.39) and CA-DNA (ρ =0.47, p=0.11) in CSF CD4+ cells. In contrast, length of time on ART was not associated with HIV-1 CA-RNA transcription (ρ =0.18, p=0.53) or HIV-1 CA-DNA in CSF cells (ρ =-0.02, p=0.95).

We failed to detect HIV-1 CA-RNA transcripts in CSF cells from two participants, both of whom had current mild HAND, and were unable to detect HIV-1 CA-DNA in CSF cells from three patients, two of whom had current mild HAND. Therefore, it was not possible to robustly analyze any association between the Double R assay biomarkers and HAND status in the current sample.

### Source of HIV-1 RNA transcripts in CSF cells

It is widely believed that monocyte/macrophage lineage cells are a major target cell for HIV-1 infection in the CNS, particularly associated with cognitive impairment (22). As the number of monocytes in the current CSF samples was so low, we instead purified monocytes from PBMC to study, using the Double R assay, whether they contain HIV-1 RNA transcripts or HIV-1 DNA. Figure 3a shows that only 6/16 (35%) of participants’ purified monocytes from PBMC had detectable HIV-1 CA-RNA transcripts and only 3/16 (18%) of participants had detectable HIV-1 CA-DNA in blood monocytes.

**Figure 3.**
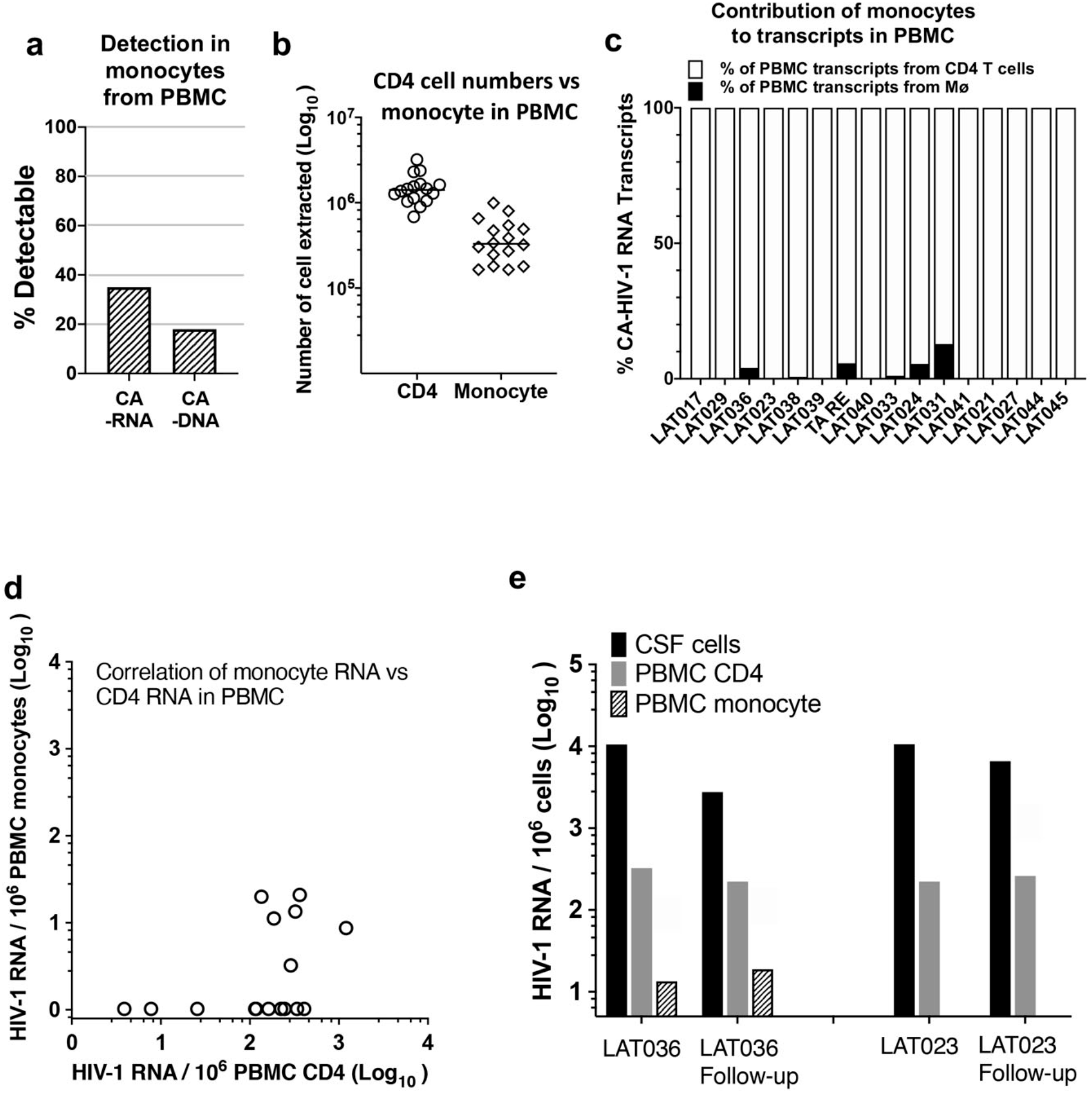
Comparison of CA HIV-1 RNA transcripts in CD4^+^T cells and monocytes in PBMCs. (**a**) Rate of detectability of cell-associated (CA) HIV-1 RNA and DNA, respectively in highly purified CD14+ monocytes from PBMC; (**b**) number of highly purified CD14+ monocytes and number of CD4+ T cells from CD14-neg PBMC used for extractions for the Double R assay; (**c**) relative contribution of CD4 T cells and CD14+ monocytes to total number of CA HIV-1 RNA transcripts from PBMC; (**d**) comparison of CA HIV-1 RNA copy numbers in CD14+ monocytes from PBMC vs CD4 T cells in PBMC, in individual patient samples; (**e**) consistent results for CA HIV-1 RNA from CSF cells, PBMC CD4 T cells and PBMC CD14+ monocytes, respectively, for longitudinal follow up samples from two participants.

The numbers of monocytes isolated from PBMC that were used to extract RNA and DNA are shown in Figure 3b. It should be noted that the number used was approximately 1,000 times higher than the number of monocytes observed in the CSF samples (Figure 1a), yet HIV-1 RNA transcripts were still not detected in 11/17 monocyte preparations from PBMC, clearly not comparable to the detectability in CSF cells. It is also important to note that we carefully monitored the number of contaminating CD4+ T cells in the monocyte preparations, with a median of 0.26% of median (IQR: 0.19-0.35%).

Furthermore, the calculated contribution of monocytes to HIV-1 CA-RNA transcripts in PBMCs is shown in Figure 3c, and is extremely low, even in the minority of cases where monocytes had detectable transcripts. The direct comparison of transcript numbers in monocytes vs those in CD4+ T cells from the same PMBC samples shows the extremely low level of HIV-1 CA-RNA transcripts in monocytes relative to CD4+ T cells, when normalized to 1 × 10^6^ cells (Figure 3d).

For 2 participants, there were collected CSF/PBMC paired samples on 2 occasions each, 6 months apart. The results show that HIV-1 CA-RNA transcripts in CSF cells, PBMC CD4+ T cells and in monocytes, respectively were maintained at very similar levels over this period (Figure 3e)

### Comparative immunological profiles of CD4+ T cells in CSF cells and in PBMCs

A very high proportion of the cells in CSF were meomry (CD45RA-negative) CD4+ T cells and expressed the chemokine receptor CXCR3 and the integrin alpha 4 (also known as CD49d), as shown in the representative flow plot, upper left of Figure 4a, but lacked the integrin ß7 (upper flow plot, second from left, Figure 4a). Furthermore, a majority of the CD4+ T cells expressed the HIV co-receptor CCR5 (upper flow plot, third from left, Figure 4a) and expressed the activation markers CD38 and HLA-DR, either singly or in combination (upper flow plot, far right, Figure 4a).

**Figure 4.**
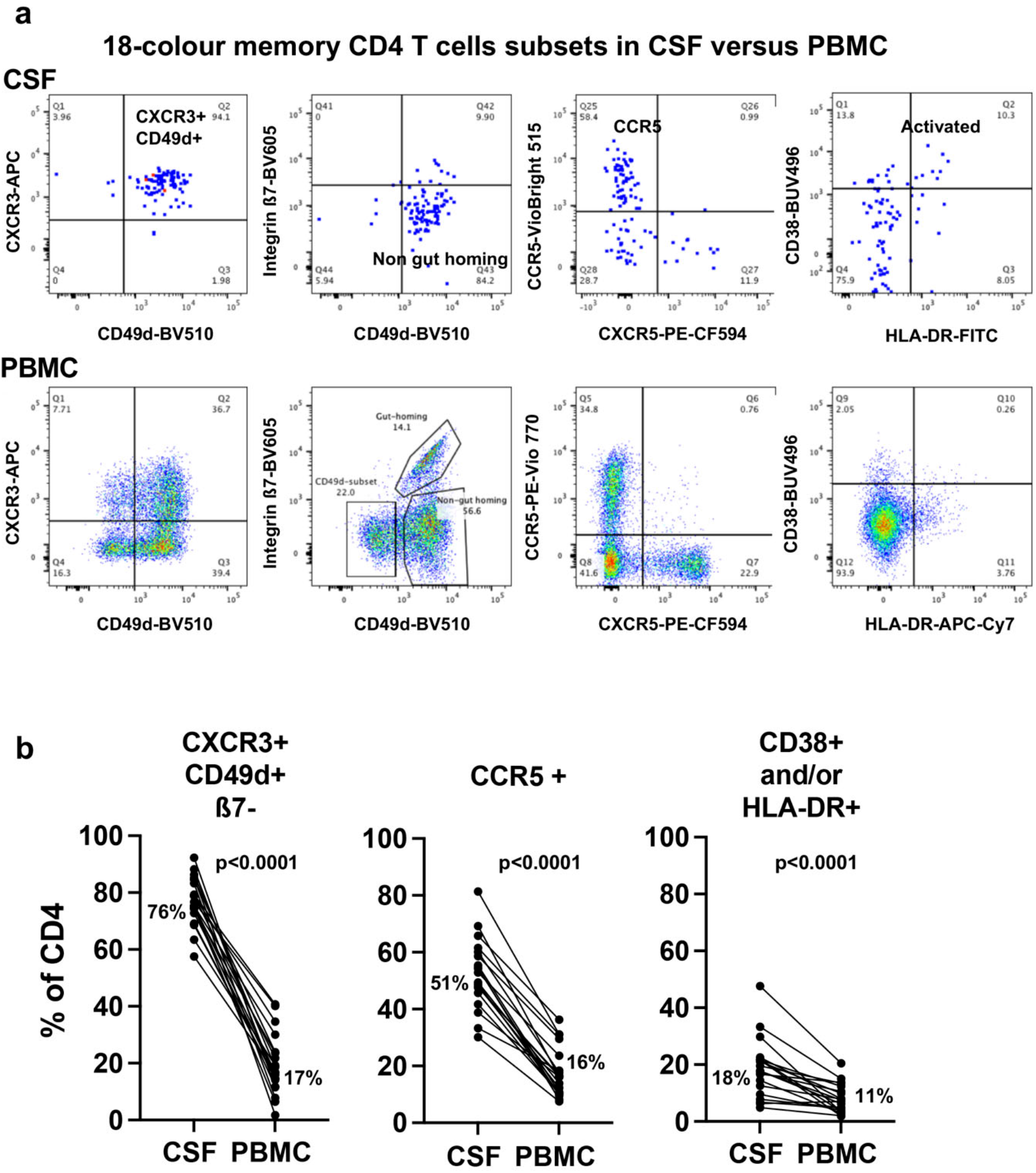
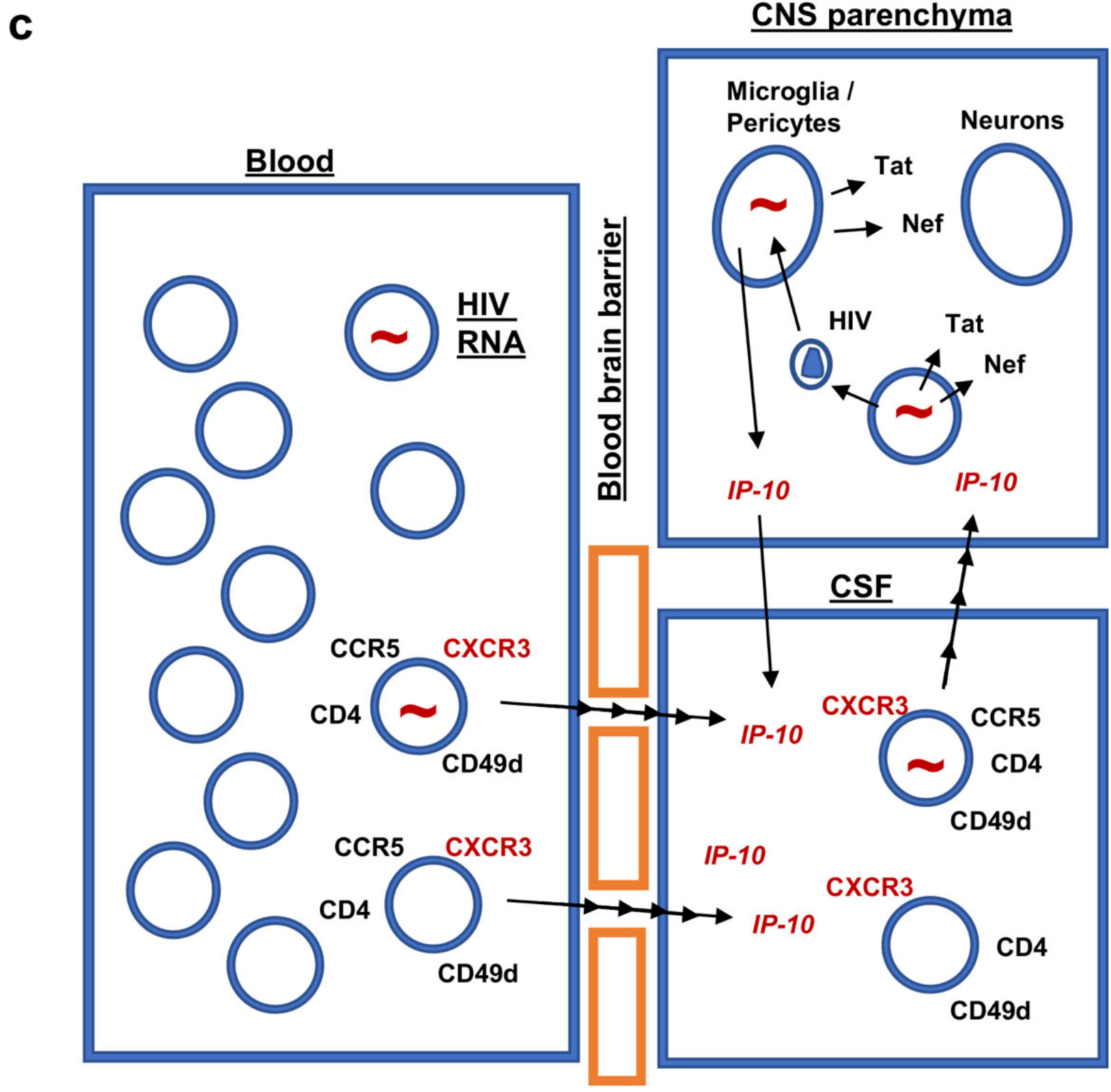
Memory CD4+ T cell subsets in CSF cells vs PBMC. (**a**) Representative flow plots showing gated memory CD4 T cells from CSF cells (upper flow plots) and PBMC (lower flow plots), respectively, stained for, from left to right, CXCR3 vs CD49d; integrin ß7 vs CD49d; CCR5 vs CXCR5; and CD38 vs HLA-DR; (**b**) comparisons of % of CXCR3+CD49d+ß7-negative cells in CD4 T cells from CSF vs PBMC; % of CCR5+ cells in CD4 T cells from CSF vs PBMC; and % of combined CD38+ and/or HLA-DR+ cells in CD4 T cells from CSF vs PBMC; and (**c**) proposed model of directed migration of CXCR3+ CD4+ T cells from blood, including HIV-infected cells, into CSF and CNS parenchyma, due to elevated concentrations of IP-10, which are in turn due to detection of HIV RNA and viral proteins by myeloid cells in brain parenchyma.

By comparison, the corresponding subsets, particularly the CXCR3+CD49d+ memory CD4+ T cells, were at a much lower proportion of CD4+ T cells in the paired PBMC samples, as shown in representative flow plots, lower row, in Figure 4a. These differences, shown in Figure 4b, were highly significant by paired Wilcoxon non-parametric comparisons (p<0.001 for the 3 comparisons, respectively; Figure 4b).

Altogether, in Figure 4c, we propose a model of trafficking of infected CXCR3+CD49d+integrin ß7-negative CCR5+ CD4+ T cells from the circulation into the CSF, as discussed in detail below.

## DISCUSSION

Using a novel, highly sensitive HIV-1 assay we found six important results: (i) the vast majority of samples of CSF cells and all samples of PBMCs contained HIV-1 CA-RNA and DNA despite undetectable cell free HIV-1 RNA by other assays of viral load, including the single copy assay; (ii) the levels per cell of HIV-1 CA-RNA and DNA were significantly greater in the CSF than those in PBMCs; (iii) CSF HIV-1 CA-RNA levels were associated with in vivo brain injury in the FWM, PCC, and to a lesser extent, the caudate area. This was driven by reduced axonal integrity (lower NAA/H_2_O) in the FWM and reduced neuronal integrity in the other brain regions; (iv) the correlation between HIV-1 CA-RNA in CSF and PBMCs was very strong; (v) the cellular composition of the CSF target cells was dominantly CD4+ T cells of the CCR5+ non-gut homing phenotype; there were very few monocytes; and (vi) the source of HIV-1 CA-RNA and DNA in CSF cells was most likely these non-gut homing CD4+ T cells; very few monocytes and other cell types were involved.

Our finding of elevated HIV-1 reservoir and its activity in CSF cells, despite undetectable cell-free HIV-1 RNA in CSF, confirm and extend recent studies. Several groups have shown that HIV-1 CA-RNA and HIV-1 CA-DNA levels were maintained at higher levels in CSF cells than in PBMCs (5, 13), but with the Double R assay we were able to detect even higher levels: 88% as opposed to 9%, for HIV-1 CA-RNA; and 76% vs 48% for CA-DNA, in a previous report from a similar cohort of patients (5). The differences in detection rates likely have two explanations. Firstly, the very low numbers of CSF cells hamper the sensitivity of standard Real-Time PCR assay or even the Droplet Digital PCR assay. Secondly, the Double R assay is much more sensitive - this is true even with adequate cell numbers as per our previous report in which we demonstrated high levels of HIV-1 CA-transcriptional activity in PBMCs despite virally suppressive ART (7).

The association between HIV-1 CA-RNA levels in CSF cells and concurrent brain injury, which was not seen with PBMC HIV-1 CA-RNA and DNA, is in keeping with the continued pathogenetic importance of HIV-1 in the CNS. Previously it has been considered that brain injury in the context of viral suppression with ART (as assessed by the absence of cell free HIV-1 in the CSF and plasma) was the consequence of pre-ART damage, intermittent low level replication, comorbidities, and other non-HIV factors (23). Our findings of very high levels of HIV-1 RNA and DNA intracellularly, make it likely that HIV-1 *still* plays a central role in neuropathogenesis. Some data have pointed to HIV-1 DNA per se in the CNS causing neuroinflammation and brain injury (24, 25). The stronger relationship of HIV-1 CA-RNA levels in CSF cells with brain injury, compared to DNA, in our study emphasizes the importance of transcription, though DNA alone may still play a role, albeit a lesser one. The nature of that transcription, however, requires further explanation. Our data relate intracellular transcriptional activity in the context of ART – what is being measured is not necessarily cell-free whole virions (as measured by standard RNA assays) but rather RNA and probably viral components before assembly into whole virions. As such, the findings indicate the presence of HIV-1 components such as Tat, Nef, Vpr, and Env, some of which are neurotoxic and immunogenic (21-24).

We defined brain injury in the current study in terms of abnormalities in ^1^H MRS of the brain and using a composite of brain major metabolites which captures the shared variance between them. This data reduction approach produces a statistically robust outcome, (17) which was ideal in the context of our sample size where too many comparisons could have led to spurious findings. Using this composite brain score injury score, we further confirm a large pool of studies which have shown that ^1^H MRS is a robust marker of brain damage in treated and virally suppressed PLHIV whether they have HAND or not (1, 26-29). This pool of research also shows that NAA and Creatine reduction (axonal/neuronal damage; reduced bioenergetics) are abnormal in PLHIV despite successful ART with viral suppression. This is in line with our finding that NAA and Creatine reduction showed a strong relationship with HIV-1 CA-RNA transcription in CSF cells.

The finding of a very strong relationship between HIV-1 CA-RNA in CSF and PBMCs is novel and suggests that HIV-1 CA-RNA is chiefly being driven from the periphery by PBMCs. Plausibly, this could occur through CD4+ T cells trafficking through the CNS with the involvement of local IP-10 production, as shown in the model in Figure 4c (and see below). However, the HIV-1 CA-RNA and CA-DNA levels in the CSF are not just a reflection of the periphery, as levels of HIV-1 RNA and DNA per CSF cell were significantly greater than those in PBMCs.

The CSF cellular origin of the HIV-1 RNA and DNA was probably the non-gut homing CXCR3+ CD49d+ integrinß7-CCR5+ memory CD4+ T cells. These were the most populous CD4+ T cell type, while there were very few monocytes. Others have also shown the CSF cells in HIV-1 infected individuals to be composed mainly of CD4+ T cells and few monocytes (30). Farhadian et al, using single cell RNA sequencing also identified microglial type cells despite only accounting for <5% of CSF cells (31). Previous studies of PBMC have shown that circulating CXCR3+ CD4+ T cells contained the highest levels of integrated HIV DNA, relative to other subsets of memory CD4+ T cells (15), and also contained the highest levels of replication competent HIV (16). Given that existing and published data confirm the dominance of CD4+ T cells in the CSF, that CD4+ T cells are the most permissive cell type for HIV-1 infection, and that CD4+ T cells carry the highest burden of HIV-1 in the blood, it is highly likely that these cells are the source of CA HIV-1 RNA transcripts in the CSF. In contrast, it appears that in the cohort in the current study, it is very unlikely that their circulating monocytes are contributing substantially to the HIV-1 RNA transcripts in the CSF cells. In support of this, a very recent study of the minority of patients on ART with CSF viral “escape”, found some evidence of the host T cell marker CD26 embedded in the envelope of cell-free HIV-1 in the CSF, consistent with a CD4+ T cell origin, rather than monocytes (32). Nevertheless, the possible roles of intermediate CD14+CD16+ monocytes, relatively rare in the circulation, but apparently enriched in the CSF cells, and microglia type cells in CSF (31), cannot be dismissed, but is still likely to be minor, based on the cell numbers.

Several aspects of our data support HIV-1 compartmentalization in the CNS, similar to previous studies (33-35). The cellular load of HIV-1 was greater in the CSF compared to blood, the cellular expression of CCR5 was greater, and perhaps most interestingly the non-gut homing CD4+ T cell phenotype was enriched in the CSF. The main chemokine ligand for CXCR3, namely IP-10 (CXLC10), has also been shown to be elevated in the CSF of untreated HIV+ patients (36, 37), correlating with CSF cell free HIV-1 RNA (36-39), leukocyte numbers in CSF (36, 38) and HAND (38). Plasma levels of IP-10 remain elevated compared to HIV-uninfected controls despite suppressive ART (40). Overall, our data strongly suggests that IP-10 is likely to lead to directed trafficking of circulating infected CXCR3+ CD49d+ ß7-negative CD4+ T cells into the CNS.

Therefore, our findings can be integrated into a model of neuropathogenesis that extends previous models into the virally suppressive ART era. CD4+ T cells, as part of normal immune surveillance, enter the CNS. Some of these cells exhibit high levels of HIV-1 RNA transcripts and are infected with replication-competent HIV-1 despite virally suppressive ART, as we have previously shown (7), and can intermittently produce low level amounts of HIV-1. This in turn leads to infection of perivascular macrophages, astrocytes, microglia and pericytes (41-43). Even those CD4+ T cells that do not have replication-competent HIV-1 still harbour high levels of HIV-1 RNA that can be translated into viral components such as Tat, Env, Nef, and Vpr that are well-described to be neurotoxic and immunogenic, (3, 42, 44-47). Of these, Tat can be detected in exosomes in CSF (9). Further, under virally suppressive ART, CNS cells are still likely to have HIV-1 RNA and DNA intracellularly which can lead to injury. This model explains why brain damage still occurs despite virally suppressive ART and why it is mild in the vast majority. ART significantly reduces the dissemination of replication-competent HIV-1 but is ineffective at inhibiting transcription, and translation of viral components, from HIV-1 integrated into the host DNA. Productive infection with whole virus formation is inhibited by ART, but not a restricted form of infection where potentially toxic viral components are still made. This model also explains the therapeutic paradox of HAND - potent antiretroviral drugs are effective against the most severe form of HAND but not against the mildest/chronic form (i.e., the current sample), as the Achilles’ heel of ART is the lack of therapy targeting HIV -1 transcription.

Our study has several limitations. First the sample size was relatively small. Nevertheless, we demonstrated robust and at least medium to large effect sizes for HIV-1 RNA transcript levels on ^l^H MRS analyses. Second, the current sample had a restricted range of cognitive performance variability. Because of this homogeneity, it was not possible to determine any association between cognitive performance and the Double R assay biomarkers. The study is ongoing and available longitudinal results suggest consistency of transcription results for CSF cells. Finally, the sample was composed only of men who were white Australians of English-speaking background. While this represents the most common demographic characteristics of the Australian HIV epidemic, further studies in more diverse samples will be important.

In conclusion, our study demonstrates high levels of HIV-1 CA-RNA transcriptional activity and DNA in CSF cells, which are associated with *in vivo* brain injury and which are likely driven by trafficking memory CD4+ T cells. These results suggest that HIV-1 still has a central role in neuropathogenesis in the ART era. Current ART needs to be extended to target inhibition of transcription from the HIV-1 promoter.

## Data Availability

All data produced in the present work are contained in the manuscript

## ACKNOWLEDGMENTS

The authors would like to thank the participants, and Positive Life NSW/NAPWHA for assisting with nationwide recruitment drive; and would like to acknowledge trial co-ordination by Fiona Kilkenny, Sarah Barney and John Ng; coordination of MRI and voxel placement for MRS by Kirsten Moffat, and PBMC biobanking by Kate Merlin, Bertha Fsadni, Sri Meka and Julie Jurczyluk.

The following reagent was obtained through the NIH HIV Reagent Program, Division of AIDS, NIAID, NIH: OM-10.1 Cells, ARP-1319, contributed by Dr. Salvatore Butera. HUT 78 Cells, ARP-89, contributed by Dr. Adi Gazdar and Dr. Robert C. Gallo.

## Author contributions

B.J.B., L.C., T.G. and K.S. conceived the project. K.S. and J.Z. designed the experiments. S.B., A.L., J.Z, S.P., and K.S. conducted experiments. C.D.R. and L.J. performed the ^l^H MRS. K.S., J.Z., Z.L., L.C. and T.G. analyzed the data and statistics. K.S., J.Z., T.G., L.C. and B.J.B. wrote the manuscript. T.I. and C.-S.H. provided the precision image pi-code (πCode) MicroDiscs detection platform. B.J.B. and L.C. provided clinical support for the study. All authors reviewed and approved the manuscript.

