## Supplementary Data and Methods for "Elevation of cell-associated HIV-1 RNA transcripts in CSF CD4+ T cells, despite suppressive antiretroviral therapy, is linked to in vivo brain injury": Suuzuki et al CSF Supplementary 23 Dec 2021.docx

**Supplementary Table 1** – Sample Demographic and Disease Characteristics

| *Demographics* |  |
| --- | --- |
| N | 16 |
| Age (years), M (SD)  Education (years), med (IQR)  Sex: male, n (%)  Ethnicity: White ESB, n (%) | 62.88 (11.84)  13 (13-15) ^a^  16 (100%)  16 (100%) |
| *HIV Disease Characteristics* |  |
| Nadir CD4+ T-Cell Count, med (IQR)  Historical AIDS, n (%)  HIV Infection Duration (years), med (IQR)  Seroconversion to ART initiation (years), med (IQR)  ART initiation > 1 year after seroconversion (n, %)  Duration on ART (years), med (IQR)  Blood CD4+ T-Cell Count, med (IQR)  Blood CD8+ T-Cell Count, med (IQR)  Past HAND, n (%)  Current HAND, n (%)  HAND Status, n (%)  GDS, med (IQR)  Mean NP T-Score, M (SD) | 185 (85-327.5)  9 (56.25%)  28 (12.75-34)  4.5 (0-10)  9 (56.25%)  22 (10.5-27)  758.5 (502.25-980.75)  701 (456.75-900.75)  6 (37.50%)  5 (33.33%) ^a^  1 (7%) ANI, 4 (27%) MND ^a^  0.4 (0-0.53) ^a^  48.31 (4.35) ^a^ |

^a^ n=15, ESB: English Speaking Background

**Supplementary Table 2** – Monoclonal antibodies used in this study

| **mAb/**  **reagent** | **Clone** | **Fluoro-chrome** | **Supplier** |
| --- | --- | --- | --- |
| (a) TruCount tube |  |  | BD |
| CD45 | 2D1 | APC-H7 | BD |
| CD56 | NCAM16.2 | APC | BD |
| CD27 | M-T271 | APC-R700 | BD |
| HLA-DR | L243 | FITC | BD |
| CD3 | SK7 | PerCP-Cy5.5 | BD |
| CD16 | 3G8 | BV421 | BD |
| CD8 | SK1 | BV510 | BD |
| CD4 | RPA-T4 | BV605 | BD |
| CD19 | SJ25C1 | BV711 | BD |
| CD123 | 7G3 | BV786 | BD |
| CD14 | MøP9 | PE | BD |
| CD38 | HB7 | PE-Cy7 | BD |
| (b) T cell phenotyping |  |  |  |
| CD28 | CD28.2 | APC | BD |
| CD27 | M-T271 | APC-R700 | BD |
| CD62L | DREG-56 | APC-eFluor780 | eBioscience |
| CXCR3 | REA232 | Biotin | Miltenyi Biotec |
| Anti-Biotin | REA746 | VioBright 515 | Miltenyi Biotec |
| CD3 | SK7 | PerCP-Cy5.5 | BD |
| CCR6 | G034E3 | BV421 | BioLegend |
| CD49d | 9F10 | BV510 | BioLegend |
| Integrin ß7 | FIB04 | BV605 | BD |
| HLA-DR | G46-6 | BV711 | BD |
| CD127 | HIL-7R-M21 | BV786 | BD |
| CCR5 | REA245 | PE | Miltenyi Biotec |
| CXCR5 | J252D4 | PE-CF594 | BioLegend |
| CD25 | M-A251 | PE-Cy5 | BD |
| CD161 | REA631 | PE-Vio770 | Miltenyi Biotec |
| CD4 | RPA-T4 | BUV395 | BD |
| CD38 | HIT2 | BUV496 | BD |
| CD45RA | HI100 | BUV 737 | BD |
| CD8a | SK1 | BUV805 | BD |

**Supplementary Table 3 –** ^1^H MRS voxel composite hierarchical linear regression models

|  | HIV-1 RNA / CSF CD4 (Log_10_) | | | |  | HIV-1 DNA / CSF CD4 (Log_10_) | | | |  | HIV-1 RNA / 10^6^ PBMC CD4 (Log_10_) | | | |  | HIV-1 DNA / 10^6^ PBMC CD4 (Log_10_) | | | |
| --- | --- | --- | --- | --- | --- | --- | --- | --- | --- | --- | --- | --- | --- | --- | --- | --- | --- | --- | --- |
| Predictors | R^2^/ΔR^2^ | B (SE) | β | p |  | R^2^/ΔR^2^ | B (SE) | β | p |  | R^2^/ΔR^2^ | B (SE) | β | p |  | R^2^/ΔR^2^ | B (SE) | β | p |
| Step 1  FWM Composite  Step 2  FWM Composite  Early/Late Treated | 0.53  0.07 | -0.26 (0.08)  -0.23 (0.08)  0.13 (0.11) | -0.73  -0.67  0.27 | 0.007  0.01  0.25 |  | 0.05  0.21 | -0.08 (0.11)  -0.04 (0.10)  0.24 (0.15) | -0.23  -0.12  0.47 | 0.47  0.69  0.14 |  | 0.16  0.37 | -0.14 (0.09)  -0.17 (0.07)  0.41 (0.14) | -0.41  -0.47  0.61 | 0.15  0.04  0.01 |  | 0.10  0.37 | -0.12 (0.11)  -0.15 (0.09)  0.46 (0.17) | -0.31  -0.38  0.61 | 0.28  0.12  0.02 |
| Step 1  PCC Composite  Step 2  PCC Composite  Early/Late Treated | 0.38  0.07 | -0.14 (0.06)  -0.13 (0.06)  0.14 (0.12) | -0.61  -0.53  0.28 | 0.03  0.06  0.29 |  | 0.09  0.19 | -0.08 (0.08)  -0.05 (0.08)  0.23 (0.15) | -0.30  -0.19  0.45 | 0.34  0.54  0.16 |  | 0.12  0.26 | -0.12 (0.09)  -0.08 (0.08)  0.34 (0.15) | -0.35    -0.23  0.52 | 0.21  0.34  0.05 |  | 0.08  0.29 | -0.11 (0.10)  -0.06 (0.09)  0.41 (0.17) | -0.28  -0.16  0.55 | 0.31  0.50  0.04 |
| Step 1  Caudate Composite  Step 2  Caudate Composite  Early/Late Treated | 0.14  0.34 | -0.14 (0.10)  -0.21 (0.09)  0.30 (0.12) | -0.38  -0.57  0.62 | 0.21  0.04  0.02 |  | 0.02  0.34 | -0.05 (0.11)  -0.13 (0.10)  0.31 (0.14) | -0.15  -0.35  0.62 | 0.65  0.24  0.06 |  | 0.02  0.41 | -0.08 (0.15)  -0.18 (0.12)  0.44 (0.15) | -0.14  -0.34  0.67 | 0.62  0.06  0.01 |  | 0.03  0.44 | -0.10 (0.17)  -0.23 (0.13)  0.51 (0.16) | -0.17  -0.37  0.69 | 0.55  0.12  0.05 |

**Supplementary Table 4 –** ^1^H MRS NAA/H_2_O hierarchical linear regression models

|  | HIV-1 RNA / CSF CD4 (Log_10_) | | | |  | HIV-1 DNA / CSF CD4 (Log_10_) | | | |  | HIV-1 RNA / 10^6^ PBMC CD4 (Log_10_) | | | |  | HIV-1 DNA / 10^6^ PBMC CD4 (Log_10_) | | | |
| --- | --- | --- | --- | --- | --- | --- | --- | --- | --- | --- | --- | --- | --- | --- | --- | --- | --- | --- | --- |
| Predictors | R^2^/ΔR^2^ | B (SE) | β | p |  | R^2^/ΔR^2^ | B (SE) | β | p |  | R^2^/ΔR^2^ | B (SE) | β | p |  | R^2^/ΔR^2^ | B (SE) | β | p |
| Step 1  FWM NAA  Step 2  FWM NAA  Early/Late Treated | 0.58  0.01 | -0.15 (0.04)  -0.14 (0.04)  0.06 (0.11) | -0.76  -0.71  0.12 | 0.004  0.01  0.61 |  | 0.07  0.18 | -0.05 (0.06)  -0.01 (0.06)  0.23 (0.16) | -0.27  -0.07  0.47 | 0.40  0.83  0.17 |  | 0.31  0.22 | -0.13 (0.06)  -0.11 (0.05)  0.32 (0.14) | -0.55  -0.47  0.48 | 0.04  0.05  0.05 |  | 0.26  0.24 | -0.13 (0.06)  -0.11 (0.06)  0.37 (0.16) | -0.51  -0.42  0.50 | 0.06  0.08  0.04 |
| Step 1  PCC NAA  Step 2  PCC NAA  Early/Late Treated | 0.29  0.09 | -0.11 (0.05)  -0.09 (0.05)  0.15 (0.13) | -0.53  -0.45  0.32 | 0.06  0.11  0.25 |  | 0.13  0.19 | -0.08 (0.07)  -0.06 (0.07)  0.22 (0.14) | -0.37  -0.27  0.44 | 0.24  0.35  0.15 |  | 0.06  0.30 | -0.07 (0.08)  -0.06 (0.06)  0.37 (0.15) | -0.25    -0.54  0.55 | 0.37  0.40  0.03 |  | 0.03  0.33 | -0.05 (0.09)  -0.04 (0.07)  0.42 (0.17) | -0.17  -0.12  0.57 | 0.56  0.62  0.03 |
| Step 1  Caudate NAA  Step 2  Caudate NAA  Early/Late Treated | 0.07  0.24 | -0.09 (0.14)  -0.12 (0.09)  0.24 (0.13) | -0.27  -0.36  0.50 | 0.37  0.21  0.09 |  | 0.06  0.31 | -0.08 (0.10)  -0.12 (0.09)  0.28 (0.13) | -0.25  -0.36  0.57 | 0.43  0.21  0.06 |  | 0.03  0.39 | -0.09 (0.13)  -0.15 (0.11)  0.42 (0.15) | -0.18  -0.32  0.64 | 0.51  0.18  0.02 |  | 0.07  0.43 | -0.14 (0.14)  -0.21 (0.11)  0.49 (0.16) | -0.26  -0.40  0.67 | 0.35  0.08  0.008 |

**Supplementary Table 5** – Neuropsychological Test Battery

| Cognitive Domain | Measure | Australian High-Functioning Male Norms  (Reference) | US Norms  (Reference) |
| --- | --- | --- | --- |
| Attention/working memory | CogState One-Back Task – Accuracy | (1) ^a^ | (1) ^a^ |
| Speed of information processing | Trail Making Test – Part A  WAIS-III Digit-Symbol Coding  CogState Detection Task – Speed  CogState Identification Task – Speed  CogState One-Back Task – Speed | Gates et al. (unpublished observations)  Gates et al. (unpublished observations)  (1) ^a^  (1) ^a^  (1) ^a^ | (2)  (3)  (1) ^a^  (1) ^a^  (1) ^a^ |
| Motor coordination | Grooved Pegboard – Dominant Hand  Grooved Pegboard – Non-Dominant Hand | Gates et al. (unpublished observations)  Gates et al. (unpublished observations) | (2) |
| New learning | Hopkins Verbal Learning Test – Revised – Total Learning  CogState One Card Learning Task – Accuracy | Gates et al. (unpublished observations)  (1) ^a^ | (4) ^b^  (1) ^a^ |
| Memory | Hopkins Verbal Learning Test – Revised – Delayed Recall | Gates et al. (unpublished observations) | (4)^b^ |
| Language | Letter Fluency – FAS  Animal Fluency | Gates et al. (unpublished observations)  Gates et al. (unpublished observations) | (2)  (2) |
| Executive functioning | Trail Making Test – Part B  Wisconsin Card Sorting Test – 64 Card Version (Computerized) – Perseverative Errors | Gates et al. (unpublished observations)  (5) ^c^ | (2)  (5) |

^a^ CogState norms are based on an Australian sample and corrected for age and gender only. Equivalent US norms are not available; Australian norms were therefore used for all participants.

^b^ US norms from the HVLT-R Professional Manual (4) are adjusted for age only. However, they were used in preference to other available demographically-corrected US norms (e.g., Norman et al. (2011) (6)) as the latter were shown to significantly underestimate performance in the Australian HIV- control sample from which the local norms were derived, whereas performance remained within expectations on the HVLT-R Professional Manual (4) US norms.

^c^ WCST-64 was not completed by the Australian HIV- control sample from which the local norms were derived; US norms were therefore used for all participants.

**Supplementary Table 6** – OM10.1 detection

|  |  | **HIV-1 DNA analysis** | **HIV-1 RNA analysis** |
| --- | --- | --- | --- |
| **Experiment setting** | **Prepared OM10 cell number** | **HIV-1 copy per 1 x 10e6 cells** | **HIV-1 copy per 1 x 10e6 cells** |
| **NO.1** | **1000** | **232** | **655** |
| **NO.2** | **200** | **50** | **114** |
| **NO.3** | **40** | **16** | **35** |
| **NO.4** | **8** | **5** | **8** |
| **NO.5** | **1.6** | **0** | **2** |
| **No.6** | **0** | **0** | **0** |

**Supplementary Table 7** – CA-HIV-1 RNA transcripts analysis with or without RT reaction step

|  |  | **HIV-1 RNA analysis with RT step** | **HIV-1 RNA analysis without RT step** |
| --- | --- | --- | --- |
| **Experiment setting** | **Prepared OM10 cell number** | **HIV-1 copy per 1 x 10e6 cells (MFI) ^a^** | **HIV-1 copy per 1 x 10e6 cells (MFI) ^a^** |
| **NO.1** | **1000** | **655 (69110)** | **0 (24)** |
| **NO.2** | **200** | **114 (50190)** | **0 (-6)** |
| **NO.3** | **40** | **35 (35442)** | **0 (63)** |
| **NO.4** | **8** | **8 (17901)** | **0 (97)** |
| **NO.5** | **1.6** | **2 (5966)** | **0 (171)** |
| **No.6** | **0** | **0 (129)** | **0 (-60)** |
| ^a^ (MFI): Mean Fluorescent Intensity, less than 200 is assay background | | | |

**Supplementary Table 8**– Evaluation of length of RT reaction time

| **ID** | **RT Reaction time (minute)** | | | | | **M** | **SD** | **CV (%)** |
| --- | --- | --- | --- | --- | --- | --- | --- | --- |
|  | **1** | **2** | **4** | **6** | **10** |  |  |  |
| **1191** | 64892 | 63295 | 67308 | 65593 | 61976 | **64613** | 2061 | **0.032** |
| **1193** | 64120 | 68888 | 65331 | 65913 | 69970 | **66844** | 2476 | **0.037** |
| **1195** | 62167 | 67529 | 71371 | 65952 | 65936 | **66591** | 3322 | **0.050** |
| Mean Fluorescent Intensity was shown. less than 200 is assay background | | | | | | | |  |
| M: Mean, SD: Standard Deviation, CV: Coefficient of Variation | | | | | | |  |  |

**Supplementary Table 9** – Evaluation of TNA extraction method for CSF cells

|  |  |  | **RNA extraction** | |  | | **TNA extraction** | | | | |
| --- | --- | --- | --- | --- | --- | --- | --- | --- | --- | --- | --- |
| **ID** | **Counted CD4 cells** |  | **RNA** | |  | | **TNA** | | **DNA** | | **RNA (TNA-DNA)** |
|  |  |  | **HIV-1 copy** | | | | **HIV-1 copy** | | **HIV-1 copy** | | **HIV-1 copy** |
| **825 Set-1** | **396,000** |  | **56.6** |  | | **150.7** | | **71.9** | | **78.8** | |
| **825 Set-2** | **99,000** |  | **32.6** |  | | **114.0** | | **28.0** | | **86.0** | |
| **825 Set-3** | **24,750** |  | **14.4** |  | | **35.9** | | **16.7** | | **19.2** | |
| **825 Set-4** | **6,188** |  | **3.5** |  | | **6.4** | | **1.7** | | **4.7** | |
| **826 Set-1** | **391,500** |  | **26.2** |  | | **103.7** | | **26.5** | | **77.2** | |
| **826 Set-2** | **97,875** |  | **9.4** |  | | **69.5** | | **11.5** | | **58.0** | |
| **826 Set-3** | **24,469** |  | **2.6** |  | | **35.1** | | **6.3** | | **28.8** | |
| **826 Set-4** | **6,117** |  | **1.5** |  | | **4.1** | | **1.1** | | **3.1** | |

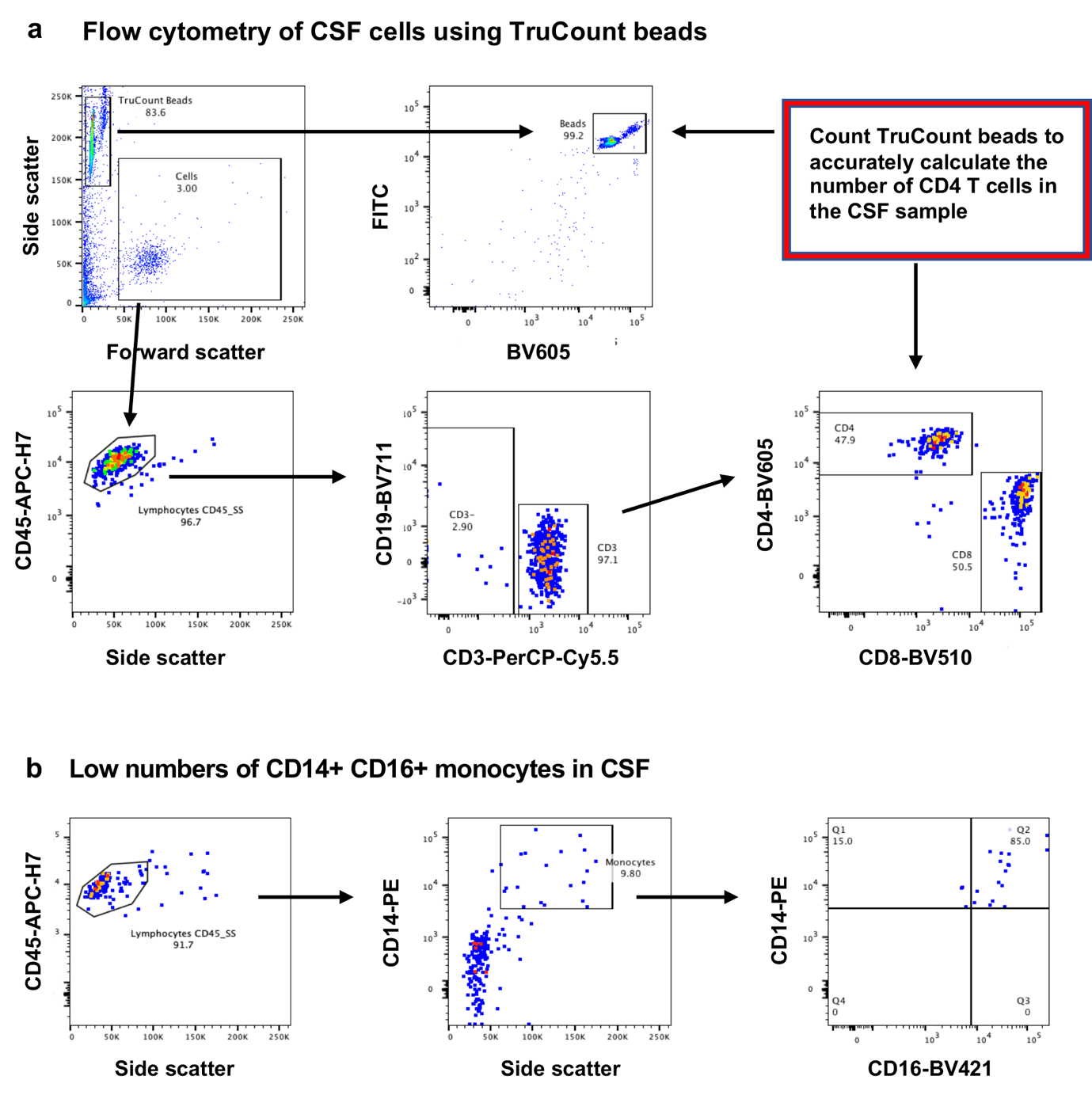

**Supplementary Figure 1.** (a) Representative flow plots showing the gating strategy to count TruCount beads and CD45+ cell types in CSF cell pellets, showing CD4+ and CD8+ T lymphocytes; and (b) the gating strategy to count CD14+ monocytes, and the presence of CD14+CD16+ intermediate monocytes in the same TruCount tubes.

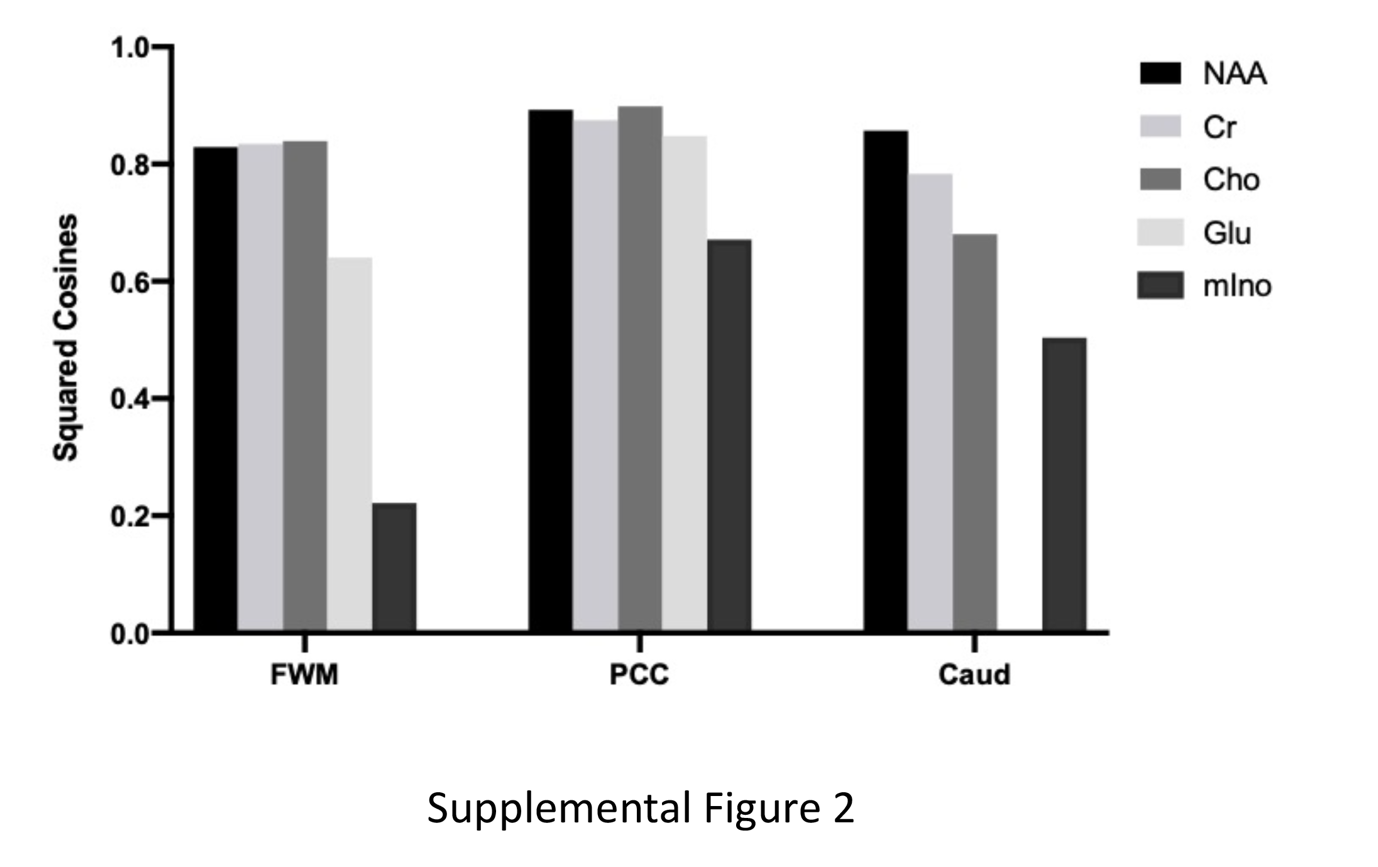

**Supplementary Figure 2**. Squared cosines of each metabolite with respect to each ^1^H MRS voxel composite score. High relative contributions of multiple metabolites reflect strong intercorrelation permitting PCA. Note the caudate composite score consists of only 4 spectra; glutamate was not able to be quantified reliably.

**Supplementary Methods**

**Neuropsychological Methods**

The standard neuropsychological battery (Supplementary Table 5) covered 7 cognitive domains, in accordance with the Frascati recommendations (7) and NeuroHIV research internationally (8), that has previously been demonstrated by our group to be sensitive to HAND (9, 10). The test battery was administered by a qualified neuropsychologist or clinical psychologist and typical completion time was around 2-3h. The test battery comprised a combination of standardized pencil-and-paper neuropsychological tests supplemented with the brief computerized battery, CogState^TM^ (11). Additionally, pre-morbid intellectual functioning was estimated using the Wechsler Test of Adult Reading (WTAR) (12) or a demographics-based regression equation when the WTAR was not administered (http://www.assessmentpsychology.com/iq-estimates-3.htm).

Psychological measures included the Depression, Anxiety, and Stress Scales (DASS-21) (13) and Mini International Psychiatric Interview (MINI v5.0.) (14). Medication adherence was assessed using the Simplified Medication Adherence Questionnaire (SMAQ) (15). Functional status was measured using a modified version of the Lawton and Brody instrumental activities of daily living (IADL) questionnaire (16).

Overall neurocognitive impairment and the presence of HAND was determined using the Global Deficit Score (GDS) method (17), a battery-wide summary of impairment. We used the standard cut-off of GDS≥0.5 to define impairment which is a well-established marker of HIV-related brain injury. The GDS was computed as follows: raw scores on each measure were converted to normally distributed and demographically-uncorrected scaled scores, and then to demographically-corrected T-scores using one of two sets of regression-based, demographically-corrected norms deemed most sensitive for detecting mild levels of cognitive impairment given the individual participant’s age, educational attainment and estimated pre-morbid full-scale IQ (FSIQ), as follows:

1) Age<67 years and either WTAR-estimated FSIQ≥110 or ≥15 years of education = Australian high-functioning male norms (Gates et al., unpublished observations). These norms were derived from a high-functioning HIV-uninfected (HIV-) MSM control sample (n=70) who participated in our NeuroHIV research program at the University of New South Wales and St Vincent’s Hospital, Sydney, Australia. The norms provided corrections for age and education in high-functioning individuals (the gender and ethnicity composition of the normative sample were homogenous, similar to the current sample. Therefore, corrections were not made for these demographic factors).

2) a) Age≥67 years; or b) Age<67 years, WTAR-estimated FSIQ<110, and <15 years of education = demographically-corrected US norms (see Supplementary Table 5).

Demographically-corrected T scores on each measure were then converted into deficit scores (ranging from 0-5), which were in turn averaged to generate the GDS.

**The Double R assay on the πCode End-Point PCR platform (OM10.1 detection)**

In order to define the assay detection sensitivity (see Supplementary Table 6), we used OM10.1 cells (containing single integrated provirus, ARP-1319, Human T-Cell Lymphoma, AIDS reagent program NIH). We made five-fold serial dilution series in the presence of one million uninfected HUT-78 cells (ARP-89, Human T-Cell Lymphoma, AIDS reagent program NIH).

No.1 setting containing 1000 OM10.1 cells in 1 x 10^6^ HUT-78 cells

No.2 setting containing 200 OM10.1 cells in 1 x 10^6^ HUT-78 cells

No.3 setting containing 40 OM10.1 cells in 1 x 10^6^ HUT-78 cells

No.4 setting containing 8 OM10.1 cells in 1 x 10^6^ HUT-78 cells

No.5 setting containing 1.6 OM10.1 cells in 1 x 10^6^ HUT-78 cells

No.6 setting containing no OM10.1 cell in 1 x 10^6^ HUT-78 cells

Two sets of identical 6-tube serial dilutions of OM10.1 and HUT-78 mixtures sample in 1ml, as above, were prepared in Eppendorf tubes, followed by centrifugation at 6000g for 5 min to form cell pellets. After removing supernatant, one set of the 6 tubes was used for CA HIV-1 RNA transcripts analysis. The other set was used for CA HIV-1 DNA analysis.

**1). CA HIV-1 RNA transcripts analysis**

RNA was extracted using the Maxwell RSC automated extraction platform (Promega) using Maxwell RSC Simply RNA Tissue kit (Cat No. AS1340, Promega) with 200μl of elution buffer. We used PrimeScript One-Step RT-PCR kit (Takara Bio, Kusatsu, Shiga, Japan), as described previously (18, 19). We adapted previously published well-described methodology for Real-Time DNA PCR for mRNA quantification (20, 21).

A DNA standard can be synthesized by cloning the target sequence into a plasmid or produced as a purified PCR product, for use in a two-step real-time PCR, with reverse transcription of sample RNA as the first step (as reviewed in (20)). DNA standards have been shown to have a larger quantification range and greater sensitivity, reproducibility, and stability than RNA standards. In contrast, the SP6/T7 in-vitro transcription method can be used to generate single strand RNA from plasmid DNA sequences cloned downstream of the SP6 or T7 promoter to generate RNA standards, but these RNA standards are not as stable as the corresponding DNA standards (20). We applied this approach for a one step Reverse Transcriptase PCR analysis (reviewed in (21)).

We did the following five critical analyses to validate the DNA standard for one-Step Reverse Transcriptase PCR analysis

1. Analysis of DNA co-extraction levels in RNA extracted solution obtained by Maxwell RSC Simply RNA Tissue kit
2. RNA integrity analysis
3. No presence of copurification of inhibitors in the RT reaction step
4. No presence of nuclease for extended storage.
5. Identification of the assay limitation factor in the RT reaction step in one step PCR reaction

**1). Absence of any DNA co-extracted with RNA using the Maxwell RSC Simply RNA Tissue kit** (see Supplementary Table 7)

Identification of DNA co-extraction levels in RNA extracted solution was essential. We carried out both analyses of PrimeScript One step RT-PCR with and without RT reaction step (Supplementary Table 7). The data indicated that DNA co-extraction levels in RNA solution were equal to zero, indicating the DNase treatment step during an automated extraction in Maxwell RSC machine succeeded in completion of removal of any DNA. HIV-1 quantification with the One-Step RT-PCR was totally dependent on the newly synthesized cDNA, generated in the initial Reverse-Transcriptase (RT) reaction at 50°C for 2min, from the positive strand of HIV-1 mRNA with the specific PCR primer. The next step DNA PCR amplification cycles (which followed the initial RT reaction in the same tube) detected only the newly synthesized cDNA from the initial RT reaction step. The quantified HIV-1 RNA data, generated by the one-step RT-PCR had no contribution of co-extracted HIV-1 DNA levels in the RNA extracted solution.

**2). RNA integrity analysis**

We assessed RNA integrity using the TapeStation analysis system (Agilent, Santa Clara, CA, USA). We confirmed that The RNA Integrity (RIN) Number was above RIN of 8 for extracted RNA using Maxwell RSC Simply RNA Tissue kit to identify generation of the best ratio of 28S and 18S rRNA peaks.

**3). No presence of copurification of inhibitors in the RT reaction step**

There are a large number of components within whole blood samples that may inhibit the RT reaction step, as well as the following DNA PCR (20, 21) for example the heme compound. We routinely use isolated PBMC, free of red blood cells, so that heme compound is not present. Nevertheless, we assessed inhibitor effect together with the RT reaction step analysis (as described below in section #5).

**4). No presence of nuclease for extended storage.**

Analysis of RNA stability is important to exclude the presence of nuclease in the extracted RNA. We assessed any possible storage effect in the RIN numbers. We found that the RIN number was maintained above RIN of 9 for extracted RNA using Maxwell RSC Simply RNA Tissue kit for 10-day storage at 4°C, indicating no presence of nuclease in the RNA extracted solution using the Maxwell RSC Simply RNA Tissue kit.

**5). Identification of the assay limitation factor in the RT reaction step in one step PCR reaction** (see Supplementary Table 8)

We found that the assay is strictly dependent on the amount of target RNA in the samples. We identified the amount of the newly synthesized cDNA levels didn’t change with extended reaction time of the initial RT reaction at 50°C: 1, 2, 4, 6, 10 minutes (see Supplementary Table 8). The data indicated that there was no difference in generated MFI (Mean Fluorescent Intensity) by the πCode End-Point PCR with extended RT reaction time. This data also suggested that there was no copurification of inhibitors in the RT reaction step.

The same data also indicates that the necessary excess amount of the specific PCR primer was present in the One-Step RT reaction. Therefore, duration of the initial RT reaction and amount of the reverse primer are not assay limitation factors in the πCode End-Point PCR. Therefore, we used 2 minutes RT reaction at 50°C, which was long enough to convert of all positive strand of HIV-1 mRNA present in the sample to quantify CA HIV-1 RNA transcripts.

Overall, the Supplementary Table 6 identified that we were able to detect CA HIV-1 RNA transcripts in less than 2 OM10.1 cells in the presence of one million of uninfected HUT-78 cells. The quantified HIV-1 RNA in five-fold serial dilution showed close to 5-fold differences in HIV-1 copy numbers in the dilution series, indicating that the πCode End-Point PCR was optimized to linearly quantify HIV-1 RNA in our developed assay system.

**1). CA HIV-1 DNA analysis**

DNA was extracted using the Maxwell RSC automated extraction platform (Promega) using Maxwell RSC Buffy Coat DNA kit (Cat No. AS1540, Promega) with 250μl of elution buffer. We used PrimeScript One step RT-PCR kit (Takara Bio, Kusatsu, Shiga, Japan) without addition of RT enzyme as described previously (18, 19). We used the HIV-1 plasmid controls: 0.73, 2.2, 6.6, 20, 59, 177, 533 and 1,600 HIV-1 copies /μl. PCR cycle conditions were identical as for our One-Step RT PCR, except omission of the initial RT reaction step. Supplementary Table 6 shows that we were able to detect CA HIV-1 DNA in 8 OM10.1 cells in the presence of one million uninfected HUT-78 cells. The quantified HIV-1 DNA in five-fold serial dilution tubes showed nearly 5-fold difference in HIV-1 copy numbers, indicating that the πCode End-Point PCR was optimized to quantify HIV-1 DNA in our system. The data in Supplementary Table 6 also indicated that OM10.1 showed very low corresponding levels of HIV-1 transcripts. The difference in copy numbers between HIV-1 DNA and CA HIV-1 RNA transcripts identified that HIV-1 RNA transcripts showed between 2-3 times higher copy numbers in HIV-1 RNA in each experimental setting of the serial dilution tubes, indicating minor ongoing transcription present in the HIV latent cells of OM10.1.

**CSF cell analysis with the Double R assay on the πCode End-Point PCR platform** (see Supplementary Table 9)

To identify HIV-1 CA HIV-1 RNA transcripts and HIV-1 DNA levels in CSF cells, we investigated methodology for small numbers of CD4^+^T cells analysis. We used the same concept as in previously described the Dried Blood Spot analyses (DBS) based on the TaqMan probe Real-Time One-Step PCR assay (below). The DBS was used as an alternative procedure instead of standard whole blood to detect HIV-1 in resource limited countries. To achieve highest detection sensitivity in DBS analysis, Total Nucleic Acid (TNA) isolation was used to recover both CA-RNA/plasma HIV-1 RNA as well as CA-DNA, where those three components of HIV-1 are present in DBS (Suzuki et al, manuscript in preparation). Furthermore, to achieve the highest sensitivity to detect HIV-1, the One-Step RT-PCR amplification was used to detect HIV-1 RNA as well as HIV-1 DNA in three automated analysis systems: Abbott M 2000, Roche Cobas Ampliprep/Cobas TaqMan, and Cepheid GeneXpert (22-25).

Although our CSF samples in this study did not contain any detectable HIV-1 RNA levels in the fluid phase, unlike in DBS samples, our CSF samples contained both CA-HIV-1 RNA and CA-HIV-1 DNA within the CSF cells. Therefore, we applied the same HIV-1 analysis approach used in DBS to adapt to CSF cell analysis. We did two critical assessments: i). selection of extraction method and ii). One-Step RT-PCR analysis for both HIV-1 RNA and DNA, to confirm the reported previous DBS analyses (22-25). We used two patients’ PBMCs, prepared using the standard Ficoll-Hypaque Plus procedure. After determining CD4^+^T cells by flow cytometry, we made three identical sets of four-fold serial dilution series in 1ml of PBMCs with Dulbecco’s Ca^2+^ and Mg^2+^ free PBS containing 2% fetal calf serum. These diluted 1ml of PBMCs in Eppendorf tubes were centrifugated at 6000g for 5 min to form cell pellets. After removing supernatant, the three identical sets were used for three Maxwell extraction methods: DNA extraction (Cat No. AS1540), RNA extraction (Cat No. AS1340), and TNA extraction (Cat No. AS1330). The accurately counted CD4^+^T cells are shown in the second column of Supplementary Table 9. The numbers of recovered CD4+ T cells from CSF samples (median: 3,605 cells), as shown in Figure 1a, therefore, the last two rows of Supplementary Table 9 are in the same range as the CSF cells, in this experiment.

The third column in the Supplementary Table 9 showed HIV-1 copy numbers by the One-Step RT PCR data by the πCode End-Point PCR with RNA extraction method. This RNA data is used as the reference in this evaluation, since RNA extraction solution with Maxwell system has no HIV-1 DNA contribution in the CA-HIV-1 RNA analysis (see above). The fourth column showed HIV-1 copy numbers by the One-Step RT PCR data with TNA extraction method. This TNA data in column four measures both CA-HIV-1 RNA and CA-HIV-1 DNA in the TNA extracted solution. The fifth column shows HIV-1 copy numbers by the DNA PCR analysis data with TNA extraction method, where we used the same PCR amplified condition as in One-Step RT RNA analysis, except without addition of RT enzyme. Therefore, the fifth column detects only HIV-1 DNA, while the sixth column shows the HIV-1 RNA copy numbers in the sample by subtraction of DNA copy number (column 5) from the total TNA HIV-1 copy number (column 4). This data shows that the calculated number of HIV-1 RNA with the TNA extraction method (column 6) showed higher levels, compared with RNA only extraction method (column 3). It is noted that especially the last two rows, comparable to CSF cell counts, showed higher levels of isolated CA-HIV-1 RNA using the TNA method. Therefore, we used 2 aliquots from the TNA extraction method with One-Step RT PCR, with and without RT respectively, and analysis by the πCode End-Point PCR, to separately determine HIV-1 CA-RNA and CA-DNA levels in our CSF cell samples.
